# Remote Photoplethysmography (rPPG): A State-of-the-Art Review

**DOI:** 10.1101/2023.10.12.23296882

**Authors:** Pireh Pirzada, Adriana Wilde, Gayle H Doherty, David Harris-Birtill

## Abstract

Peripheral oxygen saturation (*SpO*_2_) and heart rate (HR) are critical physiological measures that clinicians need to observe to decide on an emergency intervention. These measures are typically determined using a contact-based pulse oximeter. This approach may pose difficulties in many cases, such as with young children, patients with burnt or sensitive skin, cognitive impairments, and those undergoing certain medical procedures or severe illnesses. Remote Photoplethysmography (rPPG) allows for unobtrusive sensing of these vital signs in a variety of settings for health monitoring systems. Several research studies have been conducted to use rPPG for this purpose; however, there is still not a commercially available, clinically validated system that overcomes the concerns highlighted in this paper. We present a state-of-the-art review of rPPG-related research conducted including related processes and techniques, such as regions of interest (ROI) selection, extracting the raw signal, pre-processing data, applying noise reduction algorithms, Fast Fourier transforms (FFT), filtering and extracting these vital signs. Further, we present a detailed, critical evaluation of available rPPG systems. Limitations and future directions have also been identified to aid rPPG researchers in further advancing this field.

## I. Introduction

**Heart** Rate (HR) and blood oxygen saturation (*SpO*_2_) are physiological signs that are used as critical indicators of human health [1]. Also called vital signs, these typically become the first clinical alerts to a significant change in a person’s health. This is due to the existence of many complex relationships between these signs and the components of underlying circulatory and neuro-hormonal systems [1].

The first of these signs, HR, is a measure of cardiac activity, critically important in the assessment of emergent ill-health [2]. For example, a decrease in HR may be the result of an increase in intracranial pressure [3], whereas an increase may reflect hypovolemic shock [4], both medical emergencies [3], [4]. Normal values for HR in resting state lie between 60-100 BPM, with lower values being referred to as ‘bradycardia’, and higher ones, ‘tachycardia’ [5], [2].

The second of these signs, *SpO*_2_, is a broad measure of the levels of blood oxygenation, which can be defined as the percentage of Haemoglobin (Hb) in the blood carrying oxygen. When a person breathes in, oxygen enters the lungs and then transfers across the alveolar membrane and into the bloodstream [6]. The function of the circulatory system is to transfer this oxygen to all of the organs and cells of the body [7]. Once the oxygen has been off-loaded, the Hb becomes deoxygenated haemoglobin (also known as deoxy-Hb) [7], eventually returning to the heart which then sends it on to the lungs to acquire more oxygen, and so the process continues [2]. An abnormal change in *SpO*_2_ can have a serious effect on a person’s organs and is often associated with disease, typically of the lungs and/or of the heart [7]. These include pneumonia [8], [9], asthma [10], Chronic obstructive pulmonary disease (COPD) [11], [12], drug overdose [13], hypoxia [13], respiratory [14] or heart diseases [15]. A *SpO*_2_ value below 94% is considered to be abnormal for most people and indicates a significant respiratory illness, whilst some individuals with stable chronic lung disease may have a reading of 88-94% [16]. Monitoring changes in *SpO*_2_ can help identify complications including respiratory failure [14].

### A. Measuring HR and SpO_2_

Pulse oximetry is a widely used method for measuring HR and *SpO*_2_ [17], [18], [19]. It is based on the photoplethysmography (PPG) principles first introduced by Hertzman in 1937 [20]. PPG measures blood flow by calculating changes in the dispersion of light [21], [22]. This technique uses a Red and Infrared (IR) light-emitting diode (LED) and a photodetector, with a person’s finger in between. This setup allows two wavelengths of light to travel through the finger to a photodetector, which detects light not absorbed by the finger. The light absorption is governed by Beer’s law [23], whereby light is absorbed proportionately to the concentration of the light-absorbing material, such as Hb and deoxy-Hb, which means that the higher their concentration, the more light is absorbed. Lambert’s law also applies, whereby light absorbed is proportionate to the distance of the light path. This means that more light is absorbed when it travels through a longer path. As light-absorbing materials, Hb and deoxy-Hb differ: Hb absorbs more IR light than deoxy-Hb while deoxy-Hb absorbs more red light, as shown in Fig. 1. Pulse oximeter LEDs emit light at 660 nm (red) and 940 nm (Infrared) wavelengths [24]. Fig. 1 shows that at 650 nm deoxy-Hb absorbs more light than Hb, while at 940 nm the opposite is true. These physical properties are used by a pulse oximeter to measure the varying absorbance at each of the wavelengths [23].

Understanding these vitals can give important information about a person’s health and its result and performance impact clinical decisions [26]. Another form of measuring HR is via an electrocardiogram (ECG), used to characterise the cardiac rhythm in detail. However, this requires being fixed on location, typically using gel patches or chest straps [27], which might require shaving body hair. The extra adherent tape is applied to keep the electrode pad in place. This can irritate the skin and cause allergic reactions [28], [29], especially in long-term monitoring [29]. Whereas, *SpO*_2_ is currently assessed either by a simple method such as a pulse oximeter finger probe placed around the tip of the finger or by more invasive procedures such as Arterial Blood Gas (ABG) measurement where a needle is used to draw blood from an artery in the wrist [30]. Using the ABG method of getting true *SpO*_2_ can be painful and carries significant risks of infection [30], [16]. In the case of neonatal where regular sampling is required, it has a high risk of blood loss [31]. In addition to that, this process is also time-consuming [32].

**Fig. 1.**
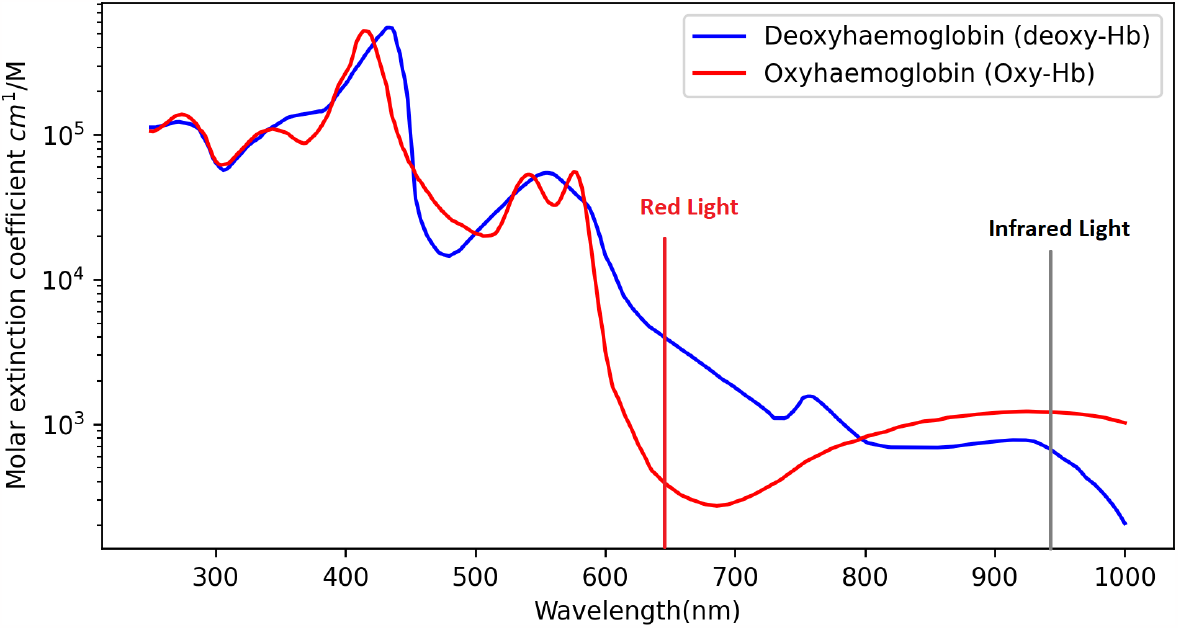
The optical extinction spectra of oxy-Hb and deoxy-Hb within the blood. Figure generated using data by Prahl [25].

A pulse oximeter device uses a clip that is mounted on a person’s fingertip^1^ or uses a strap form for example where it is wrapped on a baby’s foot^2^. Monitoring HR and *SpO*_2_ is also possible in other forms such as a wrist band^3^ [33], [34] or a smartwatch^4^, a clip that can be attached to an ear^5^ [35], or stick to the forehead^6^ [36]. Generally, a pulse oximeter clip which is mounted on a finger is the most common, accessible and cheaper form of measuring the vitals (clinical or commercial) [31], [37], [18], [19]. It is a simple and common device used in a medical setting, with relatively low intrusiveness [38], [37], [18], [19]. Recently, physicians worldwide have suggested using portable pulse oximeters for self-evaluating health vitals [39]. It plays a vital role in indicating shortness of breath related to COVID-19 [39]. It is not only widely used in clinics and hospitals [40] such as ICU but also for sports [41], [42] and within homes [43], [44], [45] due to the wide availability of commercial pulse oximeter devices. Commercial pulse oximeter devices even come with facilities that allow Bluetooth connection and their applications to enable pairing to smartphones, record vitals and store the vital history of a person [46], [47], [48], [49].

### B. Limitations of contact-based pulse oximetry

Although effective, pulse oximetry poses several problems as a monitoring tool including being open to significant error [30], [16]. The pulse oximeter clip has to be physically mounted on the finger of a person, which can be difficult to manage for adults or for patients with any form of incapacity [50] for example burnt skin [51], cognitive challenges [50], [51], during surgery [51], young children [31], sensitive skin [28] and in those with severe illness of any cause [28]. It can also be challenging to work with children or adults with physical incapacity [50]. In addition to that, it may be cumbersome to wear for long periods of time [52], restricting movement if wire-based pulse oximeter devices are used [36] and providing a constant physical reminder that their health is being monitored [28], [48].It also carries an increased risk of cross-infection [53].

Moreover, this presents a potential hazard with a high risk of choking among infants when using devices with wires [54]. Along with that, pulse oximeters have also been reported to have an increase in errors when using nail varnish [7], [50], pigmentation of skin [7], [50], access movement[7], [50], in cases where abnormal Hb and Carboxyhaemoglobin (COHb) is present and intravascular dyes; it also reduces accuracy for *SpO*_2_ that is below 83% [7]. Previous research shows that different skin pigmentation especially those with darker skin such as black or brown (i.e. with higher values in the Fitzpatrick scale^7^) had an increased error rate [57], [58]. For example, one study found a substantial bias in pulse oximeter devices used with people with darker skin pigmentation (IV-VI type from Fitzpatrick scale) had a higher error rate (5.1% ±9.2%) in comparison to white skin pigmentation (I-III type from Fitzpatrick scale) (1.9% ±10.2%) for *SpO*_2_ [59], [60]. Another study reported an increased error rate of 3.56 ±2.45% among darker skin pigmentation participants compared to white skin pigmentation participants with an error rate of 0.37 ±3.20% *SpO*_2_ [61]. Also, in a research study with anaemic participants during hypoxemia, it was observed that the error rate for the anaemic participants was 15% compared to 6.4% among non-anaemic participants [59], [40]. Different studies have been conducted where participants applied different colours of nail paint. Past studies reported conflicting results that is, some studies discovered that nail paint reduces *SpO*_2_ values [62], [63], whereas others found no effect of applying nail paint [64], [65]. One study stated that black, blue, and green nail paint decreased *SpO*_2_ reading by 3 to 6%, whereas blue nail paint decreased it from 97% to 87% [62].

### C. Remote Photoplethysmography (rPPG)

rPPG is the measurement of the flow of blood by optical means, typically involving measurement of changes in the transmission or scattering of light created by blood flow in a part of the body [54]. These changes are also reflected on the face via subtle changes on the skin where the pulse flashes lighter and darker over time. This phenomenon is not visible to the naked eye; however, it can be detected by measuring the reflectance of light over a period of time from skin pixel intensity extracted from Region of Interests (ROIs) using a camera-based system [51].

rPPG system measures the HR by analysing the skin pixel intensity of the heartbeats over time; the skin flashes darker and lighter as more and less blood flows through the region. It measures *SpO*_2_ by using the optical absorption differences across the visible and near-infrared wavelength regions between Hb and deoxy-Hb [23], [66]. Various techniques from different research systems have been evaluated, critiqued with their gaps, summarised and presented in this research paper.

## II. Research Motivation

Current contact-based methods have limitations which can be prohibitive for people with certain incapacities, severe illnesses, or burns as mentioned in section I-B which can be mitigated by developing and optimising rPPG systems. rPPG is an alternative method to measuring vital sign data such as HR and *SpO*_2_ over contact-based methods such as pulse oximetry. rPPG helps overcome the limitations of pulse oximetry which are stated in section I-B. It has the potential to revolutionise vital sign measurement by providing a more convenient and non-invasive approach to capturing physiological information from the human body without the need for physical contact. rPPG research field is expanding rapidly, but it predominantly concentrates on monitoring HR, neglecting the crucial aspect of simultaneously tracking HR and *SpO*_2_. An essential dimension of advancing these systems involves comprehending their performance in various settings, including laboratories, clinics, and homes. Unfortunately, there is currently no comprehensive review paper available that elucidates the rPPG (remote photoplethysmography) process for both HR and *SpO*_2_ and offers an in-depth analysis of the existing gaps within rPPG systems. This absence of information underscores the importance of identifying limitations and outlining future directions for researchers interested in further advancing rPPG research.

## III. Rppg Process

Previous research studies have used a variety of different equipment and set-up to capture data from participants to obtain vital signs. Thermal camera [67], [68], [69], [70], Charge-Coupled Device (CCD) camera [71], [72], other affordable web cameras or those built-in laptops [73], [74], [75], [76], [77], [78], [79], [80], [81], MS Kinect V2 [82], [83], [66], Kinect Azure [84], GoPro camera with a drone [85] and smartphone [86], [87], [88], [89], [90], [91], [92], [93], [94] cameras have been previously used to obtain a person’s vital signs. Table V and Table IV show different studies conducted with various equipment and the vital sign under observation.

The most common steps followed in an rPPG system are detailed below:

1) Face identification and ROI detection

2) Signal extraction

3) Reducing noise from signal

4) Applying Fast Fourier Transform (FFT)

5) Applying frequency filter

6) Extracting vital sign

7) Reliability check

### A. Face and ROI Detection

The first step involves identifying the person’s face from the frame. The research stationed participants in front of the camera in a static position to acquire face image frames. The participants are required to be in a specific location with no or minimal movement. In the case of minimal movement, ROIs were selected manually [95], [96], [97] whereas to cater to head movement or rotation of the head to some degree, various algorithms were used, among which Viola and Jones [98], [99] were common. One of the major reasons researchers used this library was due to its availability in the OpenCV. Researchers used a neural network-based classifier to detect ROI [100], [101], [102], a statistical model to match a person’s face to an image frame [103] and different algorithms to track features points for head movement in image frames [104]. To cater to the movement problem and identify the correct ROI, one way would be to detect face and ROI for every single frame. However, the Viola-Jones bounding box is not very accurate and can report false positives and negatives [104] along with additional computation power in the case of realtime applications [104]. Kanade-Lucas Tomasi (KLT) was used by different researchers to track feature points from a face to update ROI location [100], [105], [106], [107], [108]. Different parts of the body have been used as ROI to obtain vitals [56], [109], [110], [111], such as the entire face, forehead, and palm, as shown in Table V.

### B. Signal Extraction

Once the ROI has been identified and extracted. The next step is to extract the raw signal from the ROI image data over a specific time period. The raw signal from a colour image includes channels RGB and these values obtained from each channel of the image frame are averaged over all pixels at a time period. For example, red channel pixels averaged for ROI image data for a time period of 60 seconds (*s*_*R*_(*t*), *s*_*G*_(*t*), *s*_*B*_(*t*)) where s is the signal obtained over time t for RGB channels. This process is known as spatial pooling, and it helps average the noise contained in each pixel [51], [28], [112], [113], [114].

### C. Reducing Noise

After the raw signal has been obtained, the signal (S) containing data from all channels [*S* = *s*_*r*_, *s*_*g*_, *s*_*b*_] is passed through a Blind Source Separation (BSS), for example, Independent Component Analysis (ICA) or Principal Component Analysis (PCA) algorithm. This is applied as the signal can contain additional noise due to skin pigmentation, light source or any movement. Applying BSS helps extract a clearer signal from the raw signal input [51], [28], [112], [113], [115].

### D. FFT and filter

FFT is then applied to a raw signal to convert it into a frequency domain and apply a filter to remove unwanted frequencies. This is to increase SNR to enhance the quality of the obtained signal [116], [117]. The most common filter is a bandpass filter to remove low and high frequencies, for example, 0.65Hz and 3.5 Hz [51], [28], [112], [113], [118], [119], [120].

### E. Extracting Vital

Finally, vital sign data is to be extracted from the signal [51], [28], [112], [113], [121], [122], [123], [124], [125], [126]. A peak is identified from the signal to obtain HR vital data. Where HR is calculated by multiplying the frequency value on that peak by sixty (60) to convert it into BPM, and for Respiratory Rate (RR), it is multiplied by the ratio of the number of peaks over a time period. Researchers used different methods to obtain *SpO*_2_ from the pulse signal, which is mentioned in section V-B.

### F. Reliability Check

Only two research papers defined unique methods using SNR ratio, and previous estimates within a history window to perform consistency and reliability check to ensure history-based reliability of the vital sign data [127], [66].

## IV. Methodology

In order to identify relevant published work, we conducted a literature review. PubMed, MedlinePlus, ACM, IEEE Xplore and Springer Articles were searched using the following keywords: (“remote”, “non-contact”) and (“heart rate monitoring”, “blood oxygenation level”, “rPPG”). Literature was identified that fit the criteria detailed in Table I. The date range for the review was January 1990 – October 2022 (Figure 2). This was refined further by developing stringent exclusion criteria (also in Table I). A second round was made based using the same criteria, to include more recent research (October 2022 – September 2023). Through this round, 57 additional papers were identified.

**TABLE I.** Inclusion and Exclusion Criteria for rPPG Literature Review.

| # | Inclusion Criteria | Exclusion Criteria |
| --- | --- | --- |
| 1 | Measures HR and/or $SpO_2$ (or both) and discusses methods/techniques of measuring these vital sign data. | Articles that did not fit the inclusion criteria such as those that focused on measuring blood pressure and health monitoring such as activity monitoring.<br>Articles focused on security, perception, management, face detection and/or advantages of remote health monitoring technologies.<br>Articles focusing on economic, post-operative health monitoring involving carers.<br>Articles with incomplete or missing information regarding experiment setup, participant information and its evaluation. |
| 2 | Measures remotely using camera-based technology with participants in a conscious state within a lab, clinic or home setting. | Contact-based monitoring such as wearable (any form of pulse oximetry), wireless sensors or radars.<br>Driver state or accident monitoring. |
| 3 | Measures humans HR and/or $SpO_2$ who were able to consent for themselves. | Measures animals or rodents HR.<br>People who cannot consent for themselves. |
| 4 | Studies were published in English. | Articles that did not fit the inclusion criteria (Languages other than English). |

**Fig. 2.**
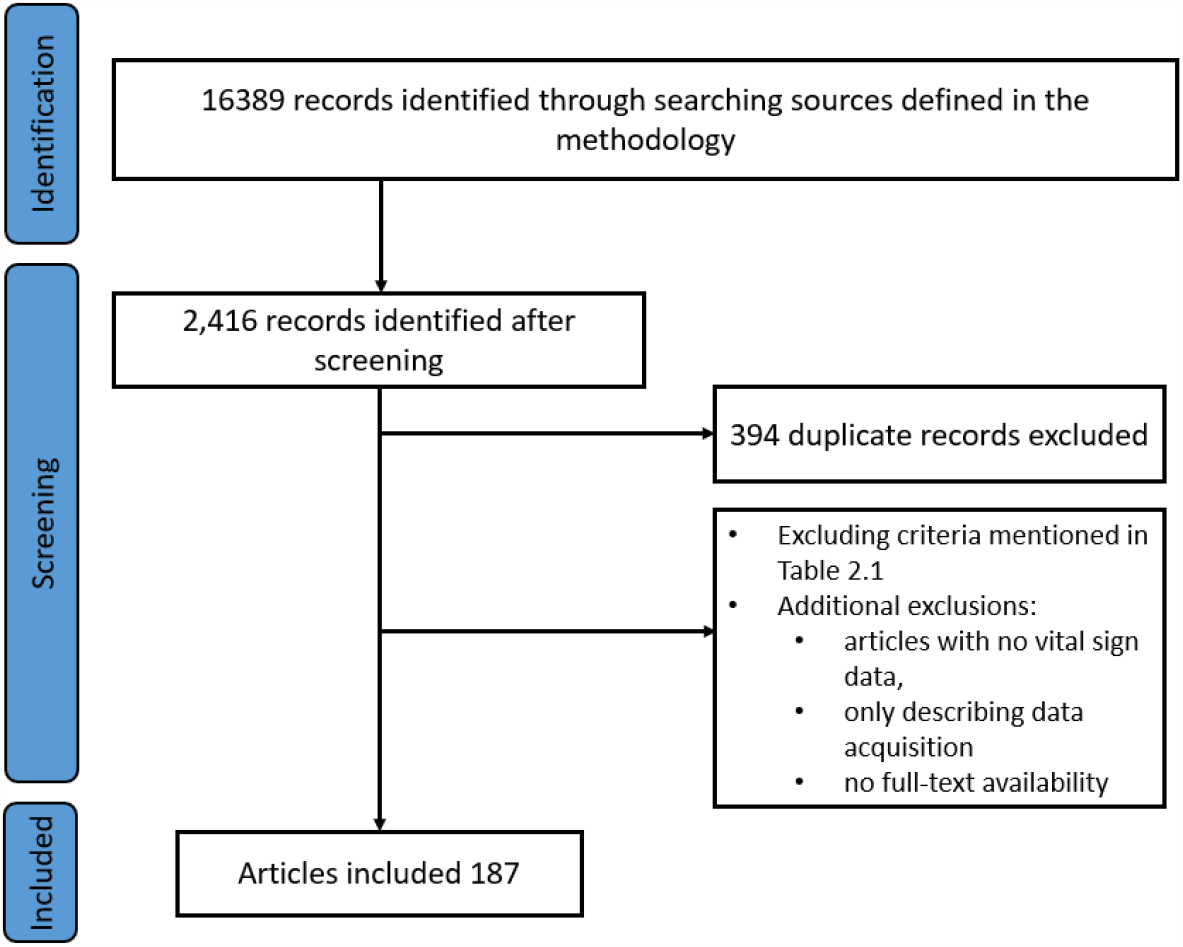
Research articles identified for the literature review based on the criteria defined in Table I.

## V. Literature Review Findings: Previous Work With Its Limitations

This section provides details of the previous research on an rPPG system, its performance, conditions, and limitations. It was found that most of the research focuses only on HR, whereas very few focus on *SpO*_2_ and only a limited number of studies focus on both. The system performance metrics (Root Mean Square Error (RMSE), r-correlation and Standard Deviation Error (*σ*)) of previous work are listed in Table III for HR and Table II for *SpO*_2_. Previous research conditions and their participant information are in Table V and Table IV.

**TABLE II.** Previous Systems evaluation measures for *SpO*_2_ performance metrics of Root mean Square error (RMSE)

| Year | Author | Participants | RMSE |
| --- | --- | --- | --- |
| 2021 | Mathew et al [86](Model1 PD) | 14 | 3.07 |
| 2021 | Mathew et al [86](Model1 PU) | 14 | 2.16 |
| 2022 | Casalino et al [128](Still) | 10 | 1.879 |
| 2022 | Casalino et al [128](talking) | 10 | 1.188 |
| 2022 | Casalino et al [128](slight rotation) | 10 | 1.881 |
| 2022 | Casalino et al [128](some rotation) | 10 | 1.063 |
| 2016 | Van Gastel et al [129] (Still) | 4 | 1.33 |
| 2016 | Van Gastel et al [129] (some movement) | 4 | 1.64 |
| 2022 | Pirzada [66] (various movement) | 40 | 2.5 |

**TABLE III.** Lab-based HR systems performance: Standard Deviation Error (*σ*), Root Mean Square Error (RMSE), and *r* correlation.

| Year | Author | $\sigma$ | RMSE | $r$ |
| --- | --- | --- | --- | --- |
| 2010 | Poh et al. [73] (Sitting still) | 2.29 | 2.29 | 0.98 |
| 2010 | Poh et al. [73](with slight movement) | 4.59 | 4.36 | 0.95 |
| 2010 | Poh et al. [73] reported by Hassan et al. [113] | 12.82 | 21.08 | 0.34 |
| 2010 | Poh et al. [73] reported by Waqar et al. [104] | 14.57 | 17.70 | 0.33 |
| 2011 | Poh et al. [74] | 0.83 | 2.29 | 1.00 |
| 2011 | Poh et al. [74] reported by Hassan et al. [113] | 12.66 | 14.01 | 0.44 |
| 2011 | Poh et al. [74] reported by Waqar et al. [104] | 18.12 | 18.02 | 0.14 |
| 2013 | Monkaresi et al. [130](ICA) | 25.54 | 35.31 | 0.53 |
| 2013 | Monkaresi et al. [130](ICA+KNN) | 4.33 | 4.33 | 0.97 |
| 2013 | Monkaresi et al. [130](ICA+KNN+Regression) | 13.70 | 13.69 | 0.58 |
| 2013 | Holton et al. [131] | 7.08 | 7.73 | 0.84 |
| 2013 | De Haan et al. [72] | 2.6 | 1.1 | 0.97 |
| 2013 | De Haan al. [72] reported by Waqar et al. [104] | 21.65 | 21.99 | -0.05 |
| 2014 | Hsu et al. [132] | - | 7.31 | 0.77 |
| 2014 | Li et al. [133] VideoHR database | 0.72 | 1.27 | 0.99 |
| 2014 | Li et al. [133] MAHNOB-HCI database | -3.30 | 7.62 | 0.81 |
| 2014 | Li et al. [133] reported by Hassan et al. [113] | 9.53 | 12.47 | 0.53 |
| 2015 | Yan et al. [134] | - | 1.2 | 0.94 |
| 2015 | Lee et al. [135] (low dynamic brightness) | - | 1.8 | - |
| 2015 | Lee et al. [135] (high dynamic brightness) | - | 4.7 | - |
| 2015 | Tran et al. [136] | - | 5.72 | - |
| 2015 | Kumar et al. [137] (still) | - | 15.74 | - |
| 2015 | Kumar et al. [137] (reading) | - | 55.34 | - |
| 2015 | Kumar et al. [137] (watching video) | - | 97.51 | - |
| 2015 | Kumar et al. [137] (talking) | - | 67.08 | - |
| 2015 | Lam et al. [138] | - | 8.9 | - |
| 2015 | Lam et al. [138] reported by Hassan et al. [113] | 8.54 | 10.34 | 0.66 |
| 2016 | Haque et al. [139] | - | 3.85 | 4.65 |
| 2016 | Haque et al. [139] reported by Hassan et al. [113] | 3.43 | 5.96 | 0.86 |
| 2017 | Al-Naji et al. [85] | - | 0.65 | - |
| 2017 | Bousefsaf et al. [140] | - | 4.81 | 0.78 |
|  |  |  | (min=2.34 and max=7.38) |  |
| 2017 | Qi et al. [141] | 3.65 | 5.0 | 0.74 |
| 2019 | Qi et al. [142] (Still) | - | 7.21 | 0.76 |
| 2019 | Qi et al. [142] (head movement) | - | 8.70 | 0.69 |
| 2019 | Qi et al. [142] (active HR) | - | 17.88 | 0.47 |
| 2021 | Wang et al. [143] | - | 8.03 | 0.27 |
| 2021 | Huang et al. [144] (MAHNOB-HCI dataset) | 6.31 | 6.42 | 0.84 |
| 2021 | Huang et al. [144] (UBFC-rPPG dataset) | 6.45 | 7.24 | 0.73 |
| 2022 | Zheng et al. [145] (low illumination) | 5.64 | 7.63 | 0.85 |
| 2022 | Zheng et al. [145] (average illumination) | 4.55 | 6.28 | 8.75 |
| 2022 | Zheng et al. [145] (high illumination) | 3.54 | 5.09 | 0.86 |
| 2022 | Zheng et al. [145] (unbalanced illumination) | 4.96 | 7.33 | 0.84 |
| 2022 | Zheng et al. [145] (slight head movement) | 5.95 | 7.03 | 0.85 |
| 2022 | Pirzada et al. [66] (various movement) | 0.018 | 7.8 | 0.85 |

**TABLE IV.**
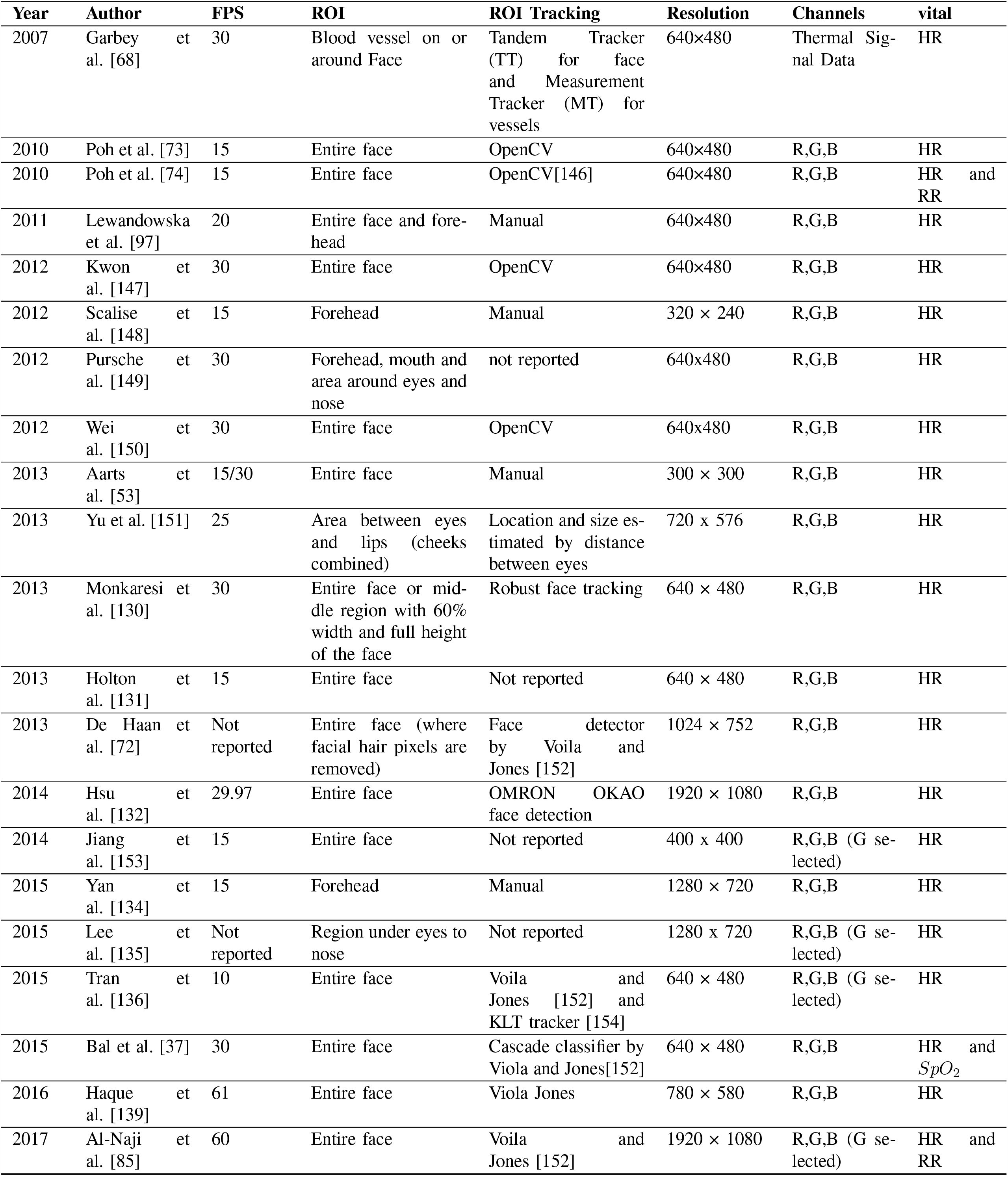
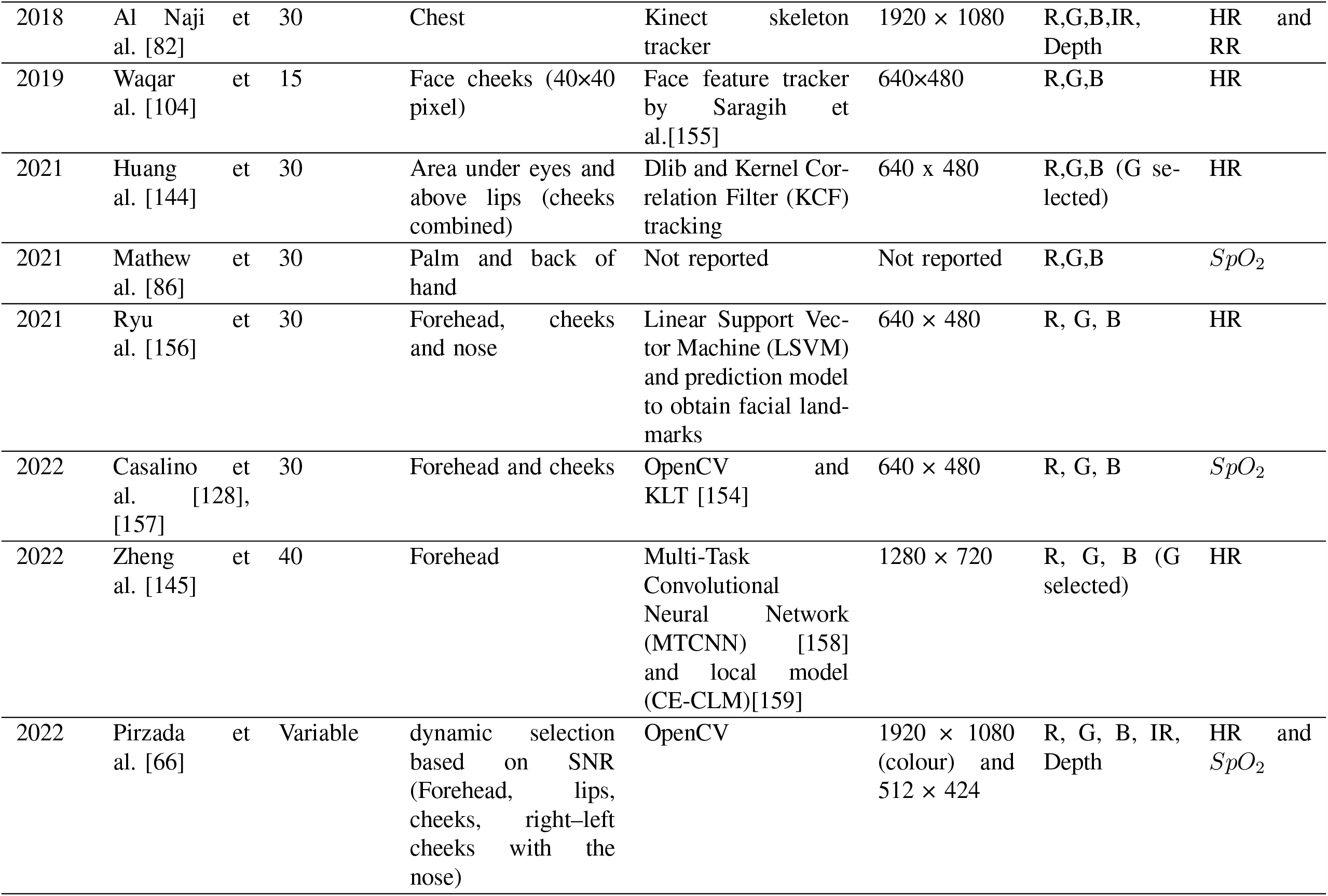
Previous Systems, FPS, ROI and Tracking detail.

**TABLE V.**
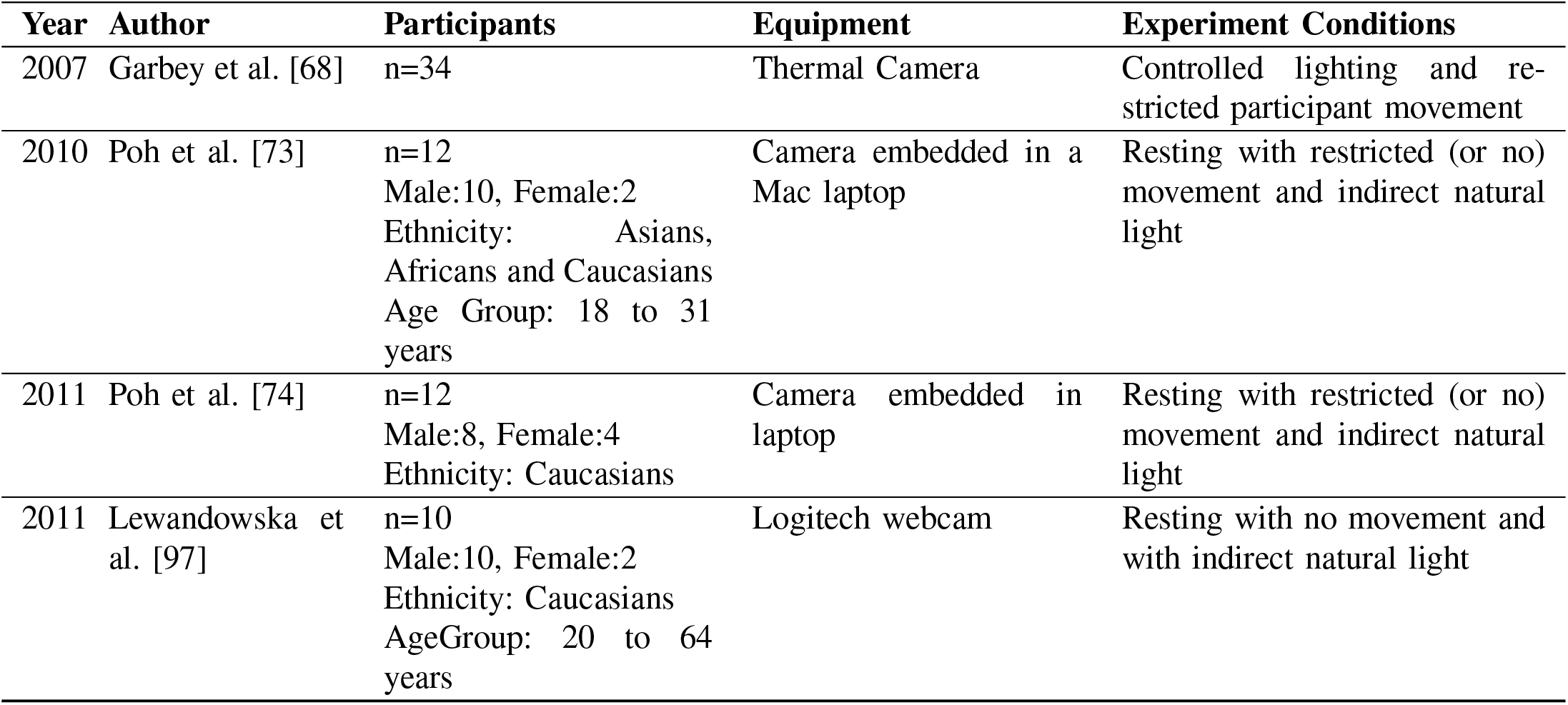
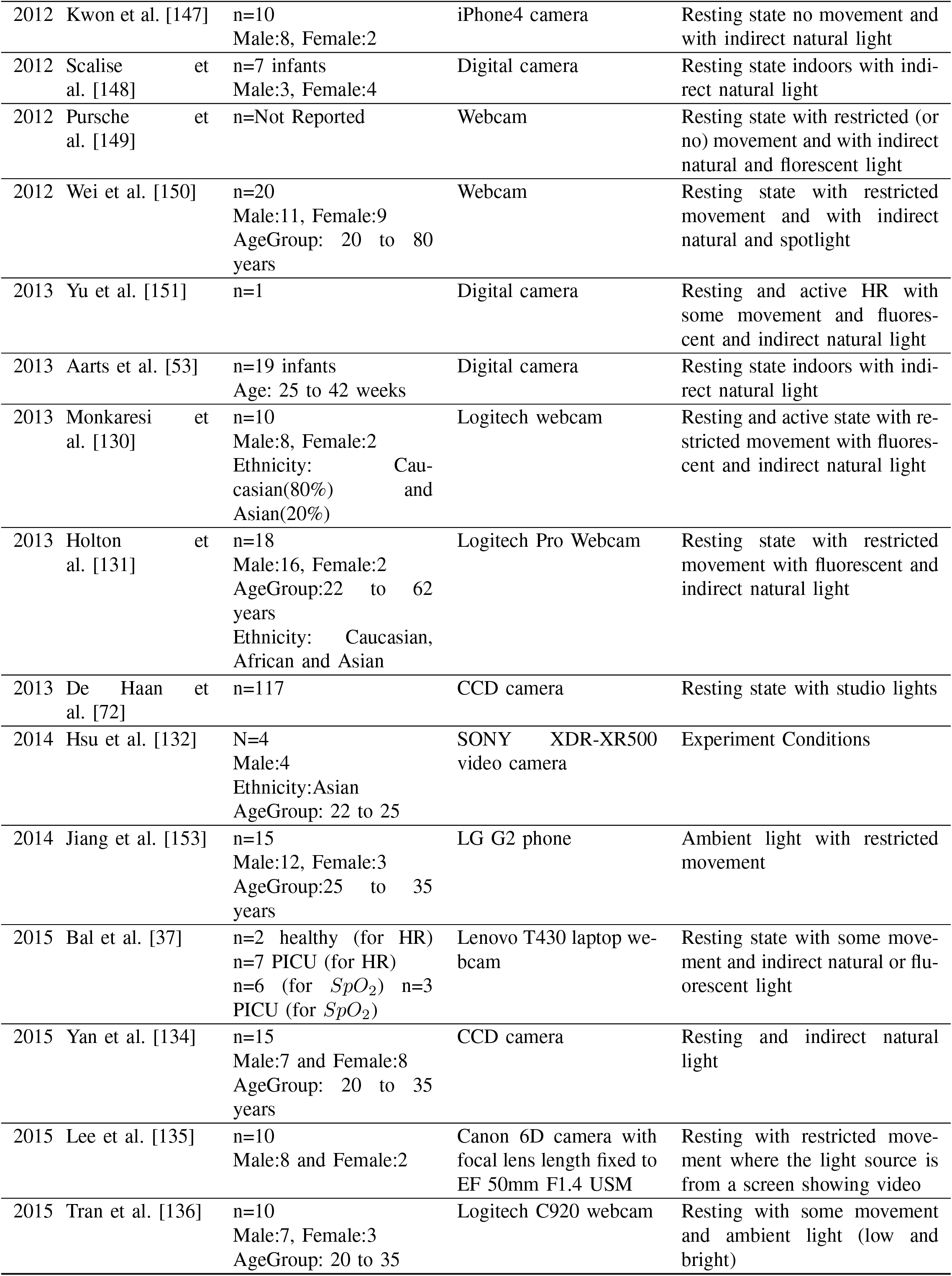
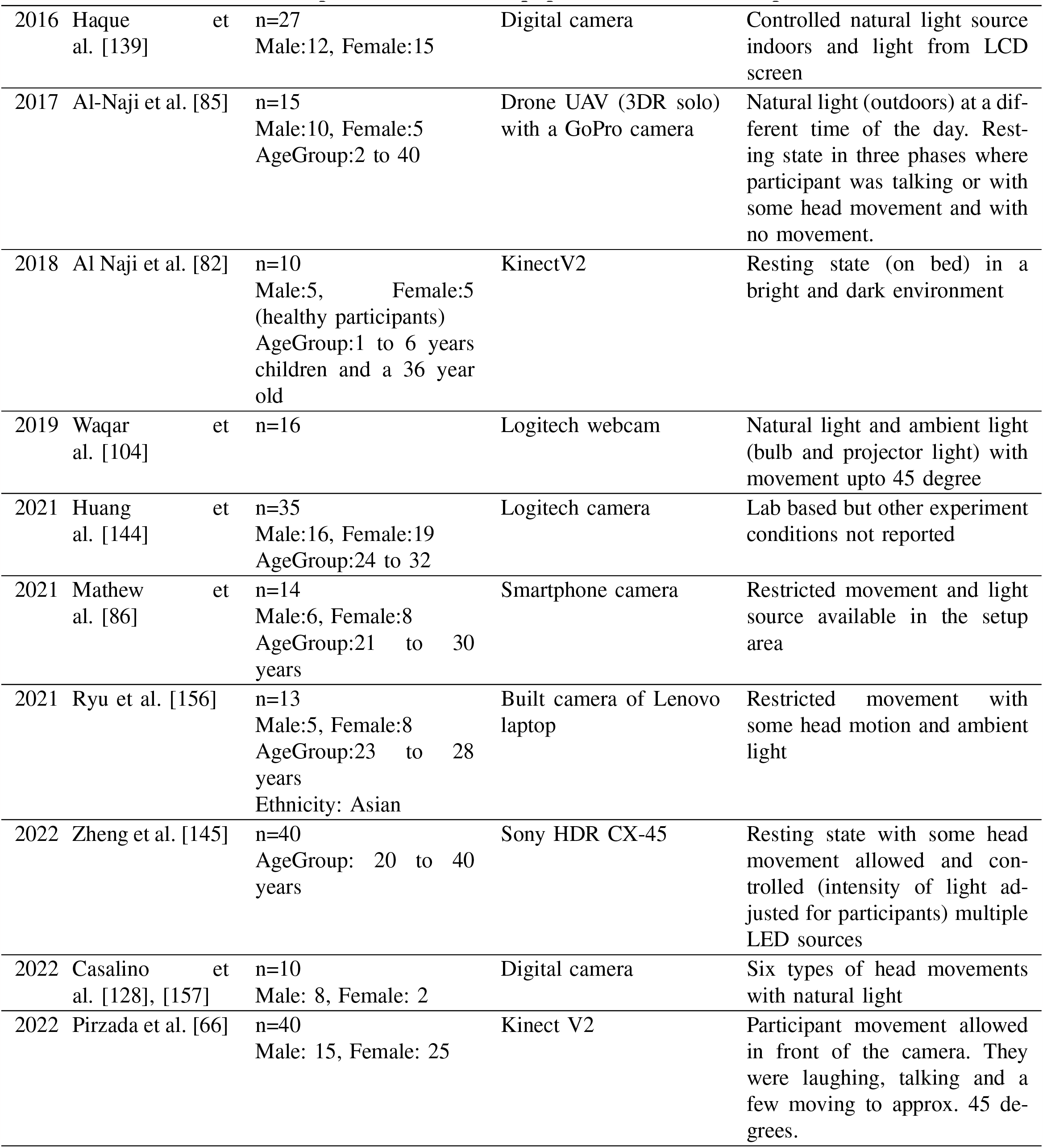
Previous Systems and Experiment Detail (n: number of participants)

### A. Remote HR Measurement

rPPG was introduced and demonstrated by using a thermal camera to record and analyse face data [67], [68]. This was to extract information about cardiac pulse, blood flow, and breathing rate. The researchers tracked the face movement and used a blood vessel (blood vessel registration) as ROI to extract a thermal signal. After extracting the thermal signal and removing noise, a Fourier Transform was applied to obtain a pulse signal. The performance of the system was reported to be 88% [67]. However, the research does not state RMSE, r-correlation or Mean Absolute Error (MAE) values. In addition to that, the research data was gathered in a controlled illumination environment where the participants’ movement was restricted (still state). The system might not perform well in a real-life scenario as illumination may vary and participants are not restricted from movement. Along with that, this thermal equipment is expensive [104], [69] which can be difficult to deploy on smaller budgets [104], [69].

Another study was also conducted utilising a thermal camera that used the forehead as ROI to measure and analyse signal data to generate HR. During data acquisition, if the participant changed the head position in any direction, the system failed to capture data from the defined ROI [69]. Researchers used the Auto-Regressive Moving Average (ARMA) model to obtain HR from the signal [160]. The biggest drawback is that even if the signal provides a reasonable result, the participant’s movement in real-life can fail the system since it only tracks the forehead. In the case of a headscarf or longer hair, it is also highly likely to produce a reading that is not accurate. Along with that, a person’s Body Temperature (BT) and environmental temperature could impact the signal extracted from a thermal video. Again, the expensive thermal equipment would also make it infeasible to deploy on a limited budget [104], [69], [161].

Remote HR measurement has also been accomplished using a CCD camera. Researchers used thirty (30) mean seconds of image data captured from the camera and analysed the change in brightness on participants’ face regions from the cheeks. After analysing the signal data obtained from participants, HR peak was identified [71]. Another research used a CCD camera to remote measure HR. It focused on participants’ movement and noise from light using chrominance rPPG to remove specular reflection variation. They used a combination of red and green channels to extract signals from defined ROI [72]. Once again, both these studies were conducted in a controlled setting which used specific lighting. Deploying this system in a real-life scenario could impact the results (inaccurate readings) due to various types of illumination present in realistic environments. CCD camera used in this research is also expensive and bulky [104], [71].

One of the early investigations to obtain HR used a laptop camera to acquire face colour images consisting of RGB channels. ICA, FFT and a frequency filter were applied to the channel data (RGB) to obtain the pulse signal. The second component (green channel) was found to typically produce the strongest pulse signal in this research study [73], [162]. This research was further refined by adding signal preprocessing such as detrending and smoothing the signal data. The system was updated to select a component with the highest peak after applying FFT and filter, which was further smoothed by moving average filter [74]. However, the study was conducted within a lab setting (in a controlled environment) with only a limited number of participants (n=12). In addition to that, restricted movement within a controlled environment does not replicate realistic environments. The face tracker used in the study detected multiple or no faces which the system handled by using the previous face captured; however, this method can fail in a real-life setting as it may not capture the real vital reading over time (using the previous face for a certain time) resulting in inaccurate HR readings.

Different researchers used various ROIs from face images obtained from participants to measure HR remotely. Face ROIs included full face, forehead, lips, eyes or nose and cheeks, which varied in pixel size. These ROIs may contain regions which cannot provide information related to HR for example eyes, teeth, beards etc., which can impact the overall performance of results. In the case, where only the mouth area is focused to obtain HR, it can impact the error rate when a person is talking or laughing as teeth do not provide a pulse signal to obtain HR. To cater to this problem, researchers [100] introduced neural networks for skin classification [163] to determine skin colour from the face image. This ROI (skin pixel) was labelled and tracked using a mean-shift tracker, which iteratively shifts points of data to the average of points of data nearby [108]. The HR was obtained after applying FFT and utilised a data adjustment scheme which resulted in obtaining HR in a short time. This work was further extended using multiple video data, which reduced the error rate [164]. However, these data do not cater to the different light sources and movement of participants and the system has not been validated in a real-life scenario.

Researchers most commonly used PCA [148], [97], [165], [166], ICA [167], [100], [168], [169], [170], [153], [171], [172], FastICA [53], [173], [151], [133], [138], [105], [174], [100], [168], [170], [153], [175], [176], RobustICA [149], Joint Approximation Diagonalization of Eigenmatrices (JADE) [147] and also Laplacian Eigenmap (LE) algorithms [150] for extracting source signals from RGB channels to obtain HR. One of the research used LE to extract signal sources from the face data obtained from participants. LE is also a dimensionality reduction technique but is nonlinear to find the internal structure of the data. The research revealed better results in comparison to other dimensionality reduction techniques. However, this has not been tested on a larger dataset obtained from realistic environments. In addition to that, the subject movement was still restricted in this research study.

A research study used Eulerian video magnification to measure changes in the face region of the participants. The data was obtained in the form of a video that was spatially decomposed into various frequency bands to which temporal filtering was applied and finally amplified by a factor. This was implemented on data where participants were in a still state [177]. Recent studies followed a similar approach to estimate HR and RR from video data as well [178], [179]. Since in real life, a person would be in motion, it is not possible to extract HR signal using this method. In addition to that, this method may not work well when there is high noise present in the signal. Research conducted also used machine learning techniques to measure remote HR [180], [181], [182], [183], [184], [185], [186]. Where the researcher [130] expanded on the work done by Poh et al. [73]. However, the accuracy drops for participants in motion and under different light sources. To counter this, the researchers [130] created a study set up to include more movement among participants. Compared to Poh et al. [73], instead of choosing a single pulse component to identify a peak, Monkaresi et al. [130] used a machine learning approach to select the pulse component among all channels (RGB components).

This approach also included applying power spectrum analysis, k-Nearest Neighbour (KNN) and linear regression to each component extracted after applying ICA to obtain features and classify them. KNN outperformed linear regression for selecting pulse signal components to obtain HR. However, generalising the model among the participants can bear different results or not perform well when new participants are introduced to the system on which it has not been trained yet and may therefore fail in a real-life scenario. Furthermore, the study was conducted in a controlled environment with specific light conditions.

To cater to challenges associated with different illumination and participant movement, Li et al. [133] suggested a technique based on Normalised Least Mean Square (NLMS) adaptive filtering and face tracking using the Viola-Jones face detector [152] to overcome these challenges. NLMS is an extension of the Least Mean Square (LMS) adaptive filter, which is used to simulate the selected filter. NLMS was used on raw signals to improve the data impacted by different light variations. To identify and locate face landmarks Discriminative Response Map Fitting (DRMF) the method was used in the initial image frame. KLT [154], [187] algorithm was then used to continue tracking the ROI. Researchers used the MAHNOB-HCI [188], [189], [190], [191] dataset for research studies to test their techniques. The results from their research exceeded the accuracy of previous studies. However, the database contains data obtained in a controlled environment with only slight movement in a lab setting. ROI tracking also fails when participants move at an extensive angle and participants’ facial expressions created noise which then shows high variations in the obtained signal. Different researchers have used datasets collected from lab setups to test and validate their methods; However, these have not been validated in realistic environments [192]. Various datasets include PURE [193], MAHNOB-HCI [194], [195], COHFACE [196], MMSE-HR [197], BH-rPPG [198], MPSC-rPPG Dataset[199] and UBFC-RPPG [200] and ARPOS dataset [66], [201].

Another study [140] suggested a method to select the most suitable ROI pixels. The researchers segmented the face region into various sub-regions on the basis of lightness dispersal. The most suitable sub-regions were then chosen and grouped by assessing their SNR [140]. Another study focused on improving signal quality by reducing noise obtained from participants’ face data by applying the block-based spatial-temporal on the face data. The spatial-temporal quality dissemination of the face ROI was then calculated depending on SNR. Adaptive ROIs were obtained by computing mean shift clustering and adaptive thresholding SNR maps, which then increased the accuracy of estimating HR [202], [203]. However, the system validation is limited to only controlled lab-based environments and needs to be stress tested for realistic environments. Another study conducted using Kinect V2 used skeleton tracking to obtain HR and RR by monitoring the chest region movement with and without a blanket over a participant (infant). This was to monitor cardiopulmonary irregularities such as bradycardia to process unclear ROI under well-lit and dark light conditions with different sleeping positions. In case of abnormalities were detected, an alarm would be triggered to notify the carer [82]. However, the system may fail if a participant is sleeping on their stomach. Another constraint is the reliability of movement features to obtain the vital, which can create inaccurate observations for different types of motions such as walking, talking or micro facial gestures. These voluntary movements can impact the reliability of the vital data obtained. A robot with a video camera incorporated has also been used to obtain HR. The aim was to design a robot for older adults to do some exercise and measure their vital [193]. However, the accuracy drops with movement and is not inexpensive which can be difficult to deploy on budget, especially for those within the middle to lower-income households around the world.

Studies conducted restrict participants from any internal or external movement. External movement includes a person moving their head or even walking around in the room, whereas internal movement is the motion of facial attributes such as talking where the mouth region is moving (moving lips) etc. In a real-life situation, these movements would be expected, which can increase noise when calculating vital signs. Research focusing on the head motion of participants from a video has been addressed by researchers [96], [204], [205], [105], [206], [207], [208], [209], [210], [211], [212], [213], [214], [215], [216], [217], [218], [219], [220], [221], [222], [223]. Where one of the researchers suggested tracking velocities of feature points on the face using Viola-Jones face detector and then tracking ROI by using KLT [96]. While another research focused only on a single ROI, specifically the forehead [224], [225], [226], this was as micro expressions below the eye line are prone to more motion as compared to forehead [225]. Researchers have also addressed this movement-based HR monitoring by using Face Quality Assessment (FQA), where low-quality image frames can be discarded, so the noise is not contributing to the final obtained values. FQA considers four parameters, including resolution, luminance, sharpness, and face trajectory, to determine if the face image requires to be removed. To track face feature points Good Feature Tracking (GFT) method was used on the image data obtained. Landmarks from the face were obtained using the Supervised Descent Method (SDM) combined with GFT to obtain face feature points and track movement and hence provided improved signal quality to obtain HR [139]. However, these motions were restricted and conducted in a controlled manner within a lab setting. The research systems were deployed by the researchers on specific hardware (labbased systems with enough RAM and Graphics Processing Unit (GPU)) to apply these techniques with controlled guided participation which does not necessarily reflect a real-life environment. People from lower to middle-income backgrounds also cannot take advantage of the system requiring expensive hardware [227], [228]. In addition to that tracking, a specific ROI such as only the forehead from the face can fail where a participant’s full face is not in front of the camera. In addition to that various research methods focusing on measuring HR need to be validated within realistic environments to check how different factors such as illumination, facial hair (such as beard), makeup and FPS impact signal data used to obtain vital sign data [229], [230], [231], [232]. To increase SNR it has been suggested to use a monochromatic camera with a green range filter. This study also used weighted average on the various face ROI. A deformable face fitting algorithm and KLT tracking [154], [233] algorithm tracked and extracted face features [137]. However, the RMSE obtained from this research is very high as mentioned in Table III which shows that it is not a feasible method to apply in a real-life system. A recent study proposed a method that uses Eulerian Video Magnification (EVM), Quality Assessment (QA) of signal, and Adaptive Chirp Model Decomposition (ACMD) to obtain HR. They validated the system in different illuminations and with some head motion [145]. Different researchers also looked into ambient, natural and varying illumination [234], [235], [236]; However, once again, the setup was within a controlled setting where participants were at a distance of 0.6m from the camera. The participant diversity information was not revealed in the research paper. In addition to that, only the forehead is selected as ROI which would fail if the participant is not facing the camera. Only one study was found that was conducted in participants’ home environments, involving different camera-to-subject distances, and shared its open dataset. However, it reported a higher error rate, particularly among individuals with darker skin pigmentation (Fitzpatrick scale IV and above) and those in an active state [66].

### B. Remote SpO_2_ Measurement

Only a few studies published presented methods to obtain *SpO*_2_. A new method to cancel aliased frequency components induced by fluorescent light flickering has previously been proposed based on autoregressive (AR) modelling and pole cancellation, which improved the effectiveness of the method under fluorescent illumination [237]. The research was conducted on patients undergoing kidney dialysis (in a resting state with minimal movement) [237]. However, movement and illumination change increased noise, in turn impacting system accuracy. The research also does not provide RMSE, r-correlation or *σ* for *SpO*_2_. While using a webcam, another research measured *SpO*_2_ and HR by using an algorithm for noise removal based on Dual-Tree Complex Wavelet Transform (DTCWT) to fix motion artefacts and artificial illumination [37].

Researchers also used R, G, and B channels to obtain *SpO*_2_ by assessing the pulse signal at two wavelengths of 660nm and 940nm [157]. This was obtained by comparing red and blue wavelength bands. However these researches [37], [157], [128], only used a 0.5m range, which is short for a realistic scenario [37], [157], [128].

The ARPOS research presented an innovative approach to measuring *SpO*_2_, by utilising extinction spectra of oxyhaemoglobin and deoxy-haemoglobin. They used a ratiometric measurement technique, dividing the recorded pixel intensity between two distinct spectral ranges, red (600–700 nm) and infrared (800–900 nm) to obtain this vital data. The error rate from this research was impressively found to be consistent over all participants for different skin pigmentation in resting and active states of ±2%. However, this needs to be clinically validated to stress test the system before this relying on this technique.

Another research used hand palms to measure *SpO*_2_ by applying spatial averaging, obtaining R, G, and B time series and applying Convolutional Neural Network (CNN) structure. However, using palms under a camera is not very practical as people in real-life would be required to keep their hands still under a camera. In addition to that, hands are also a less exposed part of the body compared to that of the face, which would make it difficult to deploy it in a real-life scenario [86], [238].

Different equipment has been used for the purpose of rPPG, which includes a CCD camera, smartphone, laptop’s webcam, drone with camera and Kinect as mentioned above. Each device has its own characteristics such as resolution, dimensions, processing power, data type collected and cost etc. Low and high-resolution digital cameras, and smartphone cameras capture RGB channels have been used in most studies to obtain vital values; whereas thermal camera allows for capturing thermal data of a participant. However, expensive equipment such as thermal [104], [69] or CCD equipment [71] (as defined in the previous section V-A) would not make it feasible to deploy on a limited budget. Smartphones need to be continuously held in hand, which can be tiring and impractical for a longer period of time or due to any incapacity.

## VI. Limitations of Existing Rppg Systems

There are several limitations in the existing rPPG research systems that researchers can focus on improving which are listed below:

### A. Equipment limitations

Thermal and CCD cameras used in these research studies can be very expensive [104], [69], [227], [228], [161], [239] to use especially for middle to lower income countries. Another problem with using a thermal camera is that it can generate temperature from the body and also the environment, which can impact the data acquired from a participant, such as thermal noise [104]. Furthermore, ambient sensing of oxygen saturation through multispectral imaging has been explored but such systems require specialist scientific equipment which can be expensive to measure *SpO*_2_ [240], [241], [242], [243], [244].

The equipment employed in these studies has primarily been limited to laboratory settings, utilising specialised hardware with ample RAM and GPU resources. However, this lab-based approach may not readily translate to real-world environments, potentially causing system failures. In clinical or home settings, individuals may be positioned at varying distances from the equipment, making it essential for the system to deliver accurate readings under diverse circumstances. This becomes especially critical in triage settings where versatile performance is essential.

Most of the research studies except the ARPOS research conducted among participants’ homes used specific hardware to acquire data where FPS from a camera was 15, 30 or 60 as shown in Table IV. This means the signal obtained from each channel (RGB) always had a constant FPS of 15, 30 or 60. The system (code) and equipment (computer and camera) were set up by the researcher in all studies and had enough processing power, RAM, graphics card and storage space to collect this data. However, people in a real-life scenario may not have access to expensive scientific equipment where the RAM, graphics card and GPU may impact the FPS and in turn, impact the number of samples in a signal obtained for each channel and if the system does not cater to this, it may impact error rate of the system. ARPOS research found that FPS impacts signal quality that is used to obtain vital data obtained data from realistic scenarios [66].

### B. Controlled Environments

rPPG studies have predominantly been conducted within controlled environments where only one study evaluated their system in participants’ homes. While it is important to initially test and validate these systems in controlled environments, it is equally crucial to evaluate their performance in realworld scenarios, particularly within clinical settings. Clinical environments encompass a wide range of heart rate and respiratory conditions, involving diverse participants. This real-world validation against clinical standards is vital for ensuring the effectiveness and reliability of such systems. Researchers need to ensure they share evaluation measures when discussing the results of their research systems to ensure further progression in the rPPG field.

Currently, there is no rPPG system that is currently commercially available that can monitor these vital signs remotely unobtrusively and has been clinically validated. It is crucial for such systems to be validated in real-life environments including labs, homes and clinics to stress these systems. Furthermore, the effectiveness of these systems for monitoring health in clinical settings has yet to be fully validated. It is crucial that rPPG systems demonstrate the ability to promptly identify patients at the highest risk of health deterioration, highlighting the significant advantages of utilising such remote technology in healthcare.

### C. ROI Selection and Occlusion

Moreover, relying solely on a single Region of Interest (ROI), like the forehead or cheeks, can prove ineffective in measuring vital signs. This limitation becomes evident when the chosen ROI is obscured from the camera’s view, for instance, when a person wears a headscarf or has long hair that conceals the forehead, or when the individual is positioned at an oblique angle to the camera. Therefore, it is crucial to subject a system to comprehensive testing across diverse settings to validate its performance and evaluate its error rate. Tracking people can also be occluded by objects, other people or both from the camera’s view to send data to the system which is common for front-viewing cameras utilised by the systems [245]. Due to occlusion, a user’s skeleton cannot be identified [245]; however, existing research has identified several methods to help solve this issue which includes API tracking of multiple people’s positions in an environment where occlusion occurs [246], [247], [245], location-aware wearable haptics [248], sound localisation [249], using API, a toolkit to track and multiple cameras within an environment to track multiple peoples and joints using depth data [245] and using shadow and skeletal fusion data to track people within an environment [250]. Occlusion needs to be further studied related to the rPPG systems.

### D. Participant Diversity and Experiment Conditions

Most of the research mentioned in Table V and Table II have a limited number of participants with a narrow range of skin pigmentations. Specifically, when examining studies related to *SpO*_2_, only one of them featured a substantial participant group of 40 [66], while the remaining studies included fewer than 15 participants, as indicated in Table II. For HR measurement, a similar trend emerges with most studies having a limited number of participants. Only three studies stood out with a larger number of participants [145], [66], [72]. Among these, two studies involved 40 participants each [145], [66]. However, only one of these studies collected vital sign data within realistic home environments, encompassing both HR and *SpO*_2_ measurements [66]. The third study [72], which included a substantial number of participants of 117, only focused on measuring HR and was conducted in a controlled environment resembling a studio setup with limited movement and utilising expensive equipment [72]. Whereas the rest of the research studies in the Table V had a much lower number of participants with restricted experimented conditions. Furthermore, an important aspect is that, most of the studies neglected to measure oxygenation levels at a distance, as shown in Table IV.

All the research considered only one participant at a time except a few studies which included 2, 3 or even 6 participants measuring at a time [51], [66]. Measuring multiple participants simultaneously is still a feature required, especially when multiple people are within an environment. All the research studies mentioned above had participants closer to the camera (up to a distance of 0.5 from the camera) and only two studies considered measuring HR up to 4m [82], [66]. Most of the studies also do not disclose participant characteristics. A wider range of demographics such as various skin pigmentation and ethnicity need to be included in the research studies so it’s not only validated and made available for one specific group of people but can benefit everyone.

In a real-time situation, participants’ environments may vary such as triage environments in clinics, care homes or people’s homes and different factors such as face rotation, facial expression, varied distance from the camera, illumination, beards, skin pigmentation etc. can be present which can impact the system’s obtained data. Parameters related to makeup and beard were only limited to two studies, however with only a small number of participants [136], [66] and one study also does not analyse or segment results based on these parameters [136]. These experiments are important to be conducted within different age groups, skin pigmentation and different environmental parameters.

### E. Dataset

Most of the studies did not share open data and code from their research. Only a limited number of public databases are available; however, they also are conducted within a lab environment with restricted parameters such as light, movement and not realistic distance from the camera. There is a lack of publicly available data sets collected from a reallife environment, so the system evaluation measures can be tested on those to evaluate their system [51], [66]. Only one dataset was found collected from the home environments of participants but this does not include participants with black skin pigmentation [201]. Studies conducted restrict participants from micro facial gestures or physical movement during vital data acquisition. In addition to that, if the error rate is not evaluated for realistic environments with natural movement or for example during undergoing treatment, it can result in failure to accurately measure vitals due to the presence of noise. Therefore, it is essential to measure vital sign data in real-life environments [51], [66]. Face tracking will also impact ROI selection when movement takes place, depending upon the lighting, and exposure length of the camera, it could potentially create blurry frames, and it is important to analyse data with noise to ensure the system will be able to perform as required in a real-life environment [51]. It would be more beneficial if video streams of Colour, IR and Depth Videos would be made available open source so researchers can analyse the realistic data including various movements, illumination, environments, skin pigmentation and other factors with their respective timestamps to analyse their algorithms.

## VII. Bias in Rppg Systems

Discussing rPPG systems presents several challenges, particularly because they rely on camera-based technology focused on capturing facial data. These challenges become especially noticeable due to the diversity of participants’ skin pigmentation. rPPG systems must be accurate, as they fall under medical technology upon which people may depend once they become commercially available. Therefore, rigorous validation and extensive testing across a diverse range of participants are imperative. Many publicly available databases primarily emphasise individuals with lighter skin pigmentation, typically categorised under the Fitzpatrick scale I-III. Neglecting diversity can pose a significant disadvantage when deploying rPPG systems; making it beneficial only for a specific group of people. One study [137], as shown in Table III, demonstrated a higher error rate in such cases. In contrast, a recent study, ARPOS [66], showcased a lower error rate and introduced effective techniques across white and darker skin pigmentations. However, the study’s dataset did not include any black participants.

Another critical observation from this review is the limited consideration given to participants who wear makeup, such as lipsticks or concealers, or those with facial hair, ranging from light to heavy beards. These factors can significantly influence the accuracy of data readings in rPPG systems. Only two studies were found which stated participants wearing makeup[66], [251]. Both research studies found makeup in Colour and IR impacted the accuracy of the rPPG systems and suggested further rigours testing. However, the ARPOS study only had one participant wearing lipstick [66] whereas the other research study only had participants wearing foundation [251]. Consequently, it becomes important to rigorously validate these systems, accounting for these various biases, and further evaluating their performance across different genders and age groups.

## VIII. Summary

This paper presents an in-depth literature review on rPPG-related research that can measure HR and/or *SpO*_2_ of people at a distance. Various processes, techniques, participant characteristics, equipment and experiment conditions used in the research studies were discussed. The performance and evaluation measures of previous research and its limitations were presented in this paper as well. Various equipment that was utilised included a thermal, CCD, web camera, Kinect V2, and camera installed in robots, drones and smartphone cameras have been detailed in this paper. The common process for extracting these vital signs usually involves ROI selection, extracting raw signals, preprocessing data, applying algorithms, FFT, filtering and identifying vitals. The review found that most research studies focused on measuring HR, a few on *SpO*_2_ and only a very limited number of research focused on measuring both vital sign data in a remote manner.

Most of the research studies for measuring HR had low participants and only a couple had a higher number of participants (as shown in Table V and discussed in section VI-D); but were conducted in a very controlled setup or lab environments which cannot be replicated in real-life scenarios. In addition to research data being gathered mostly in a controlled environment or a lab setting; it also restricted participants from micro facial gestures or physical movement of their arms or face rotations as in a real-life situation people cannot be expected to be very still for multiple minutes when their vitals are being measured as natural movement is normal (such as talking, rotation and movement due to facial expressions) which can increase noise in the data.

There is only one real-life scenario database publicly available collected to validate rPPG systems [66]. Previous research needs to consider various factors when designing an rPPG system including varying camera-to-subject distances, fluctuating frames per second (FPS), computational resource requirements, diverse environments, illumination conditions and participant diversity. Furthermore, it is crucial to validate and test for performance under conditions involving natural movements during vital sign measurements that may lead to inaccurate results. This issue becomes particularly critical when considering real-world deployments such as clinics and hospitals.

In conclusion, the field of rPPG has made significant progress in recent years. This review has highlighted various aspects of these methods for measuring HR and *SpO*_2_. Several important findings and research challenges were been identified. rPPG has enormous potential for various applications in both clinical and non-clinical settings. However, addressing the identified research challenges is crucial for advancing this field and ensuring the accuracy and reliability of these systems in real-world scenarios. This review serves as a valuable resource for researchers seeking to explore and contribute to this evolving field.

## Data Availability

All data produced in the present work are contained in the manuscript.

## IX. Acknowledgments

In order to meet institutional and research funder open access requirements, any accepted manuscript arising shall be open access under a Creative Commons Attribution (CC BY) reuse licence with zero embargo.

## Footnotes

1 Finger pulse oximeter (Accessed 25/4/2023), https://bit.ly/3KqORCo

2 Baby’s foot pulse oximeter (Accessed 25/4/2023), https://bit.ly/3y4duCu

3 Wrist pulse oximeter (Accessed 25/04/2023), https://bit.ly/3rXsDlp

4 AppleWatch6 (Accessed 25/4/2023), https://www.apple.com/uk/apple-watch-se/

5 Ear pulse oximeter (Accessed 25/4/2023), https://bit.ly/38z4ASH

6 Forehead pulse oximeter (Accessed 25/04/2023), https://bit.ly/3vn0KFq

7 The Fitzpatrick Scale ranges from I (epidermal melanin around 3% volume as the lowest in scale) to VI (epidermal melanin 43% volume as the highest in scale)[55]. Type I-III: Pale to Fair White skin pigmentation, Type IV-V: Light and Dark Brown skin pigmentation, and Type VI: Black skin pigmentation[56].

